# Generalized Sensory Sensitivity for Prediction of Post-Surgical Analgesic Outcomes: An Observational Cohort Study of Total Hip Arthroplasty and Hysterectomy

**DOI:** 10.64898/2026.05.26.26354108

**Authors:** Andrew Schrepf, Tristin Smith, Noah Waller, Richard E. Harris, Eric Ichesco, Chelsea M. Kaplan, Sara R. Till, David A. Williams, Sawsan As-Sanie, Julia M. Evanski, Andrew Urquhart, Chad M. Brummett, Daniel J. Clauw, Steven E. Harte

## Abstract

**Background:** A substantial minority (∼20%) of patients fail to achieve meaningful pain reduction following surgery intended to relieve pain. Risk is elevated in patients with nociplastic pain features, but available self-report measures were not designed for pre-surgical screening. We aimed to develop a brief, data-driven screener for poor analgesic response to surgery.

**Methods:** Participants were recruited from tertiary orthopedic and chronic pelvic pain clinics. Total hip arthroplasty participants had Kellgren–Lawrence grades III–IV with hip pain ≥1 year; hysterectomy participants had chronic pelvic pain ≥6 months. The primary outcome was a 50% reduction in worst pain at six months. Items were selected via elastic net regression with k-fold cross-validation from 68 candidates.

**Results:** Of 428 participants (81% female; mean age 51), 35% failed to achieve a 50% pain reduction. The resulting 11-item screener — the GenerAlized sensory sensitivity for sUrGical rEsponsiveness (GAUGE) — comprises pain across seven body regions and four symptom items measuring interoception (nausea, numbness/tingling) and exteroception (sensitivity to sound, sensitivity to odors). GAUGE outperformed the Central Sensitization Inventory, Fibromyalgia Survey Criteria, and PainDETECT for predicting surgical non-response (RR 1.535, 95% CI 1.342–1.55; AUC 0.738; sensitivity 0.741, specificity 0.635) and for predicting Patient Global Impression of Change. In an independent validation cohort of 54 total knee arthroplasty patients, GAUGE outperformed the Fibromyalgia Survey Criteria in predicting pain severity at six-months.

**Conclusions:** GAUGE is a data-driven, theoretically grounded screener for poor analgesic response to surgery, with potential utility for pre-surgical counseling and clinical trial enrichment.

## INTRODUCTION

More than 600,000 total hip arthroplasty (THA) procedures and more than 500,000 hysterectomies are performed annually in the United States, for which chronic pain is a common indication.^1, 2^ Unfortunately, 20% or more of individuals will not experience pain relief after surgery, leading to considerable dissatisfaction for both patients and providers, in addition to the economic burden of failed treatment.^3, 4^ When the removal of damaged tissues and associated inflammatory pain-generating signals does not improve pain, the implication is that central nervous system (CNS) sensitization may be responsible, but there is no clinical guidance available for assessing CNS sensitization prior to surgery. A handful of promising studies show that elevated scores on self-report measures assessing symptoms associated with CNS sensitization, (e.g., widespread pain, sensory sensitivity, negative mood) predict poorer analgesic responses to surgery.^5–7^ Identifying patients at risk for surgical non-responsiveness prior to the procedure would improve patient counseling, promote the development of adjuvant or alternative treatments, reduce costs, and lead to greater patient satisfaction and quality of life. Available measures, (e.g., Central Sensitization Inventory^8^, Fibromyalgia Survey Criteria^9–11^), however, have several notable drawbacks that diminish their utility in a preoperative setting.

A useful screening tool should be brief and constructed from individual items that are highly predictive of poor analgesic outcomes after surgery. Here we formally evaluate the predictive power of self-report symptoms from domains associated with CNS sensitization for the purpose of constructing a brief screening measure for analgesic non-response to surgery. Using two prospectively collected surgical cohorts undergoing procedures to relieve chronic pain (i.e., THA, hysterectomy,), we evaluate the strength of individual self-report items using cross-validation techniques, and then formally test the performance of this new measure against the Fibromyalgia Survey Criteria, Central Sensitization Inventory, and painDETECT.^8–11^ The result is a new, brief self-report instrument, the GAUGE (GenerAlized sensory sensitivity for sUrGical rEsponsiveness) screener that outperforms currently available measures.

## METHODS

### Sample

Participants were drawn from parallel studies conducted at Michigan Medicine (University of Michigan, Ann Arbor, MI) which shared protocols for the investigation of surgical analgesic non-response. These included individuals with hip osteoarthritis scheduled to undergo THA from the Mechanisms of the Centralized Pain Phenotype study (MiCAPP; HUM00120181), and individuals with chronic pelvic pain scheduled for hysterectomy from the Peripheral and Central Nervous System Mechanisms of Persistent Post-Hysterectomy Pain study (MiHYST; HUM00117473). The studies were conducted between 2016-2023. A Consort diagram showing numbers of individuals approached to the final analyzed sample is shown in **Figure 1**. Summary inclusion and exclusion criteria are presented in **Figure 2** for the overall sample and each cohort alongside the study design.

**Figure 1.**
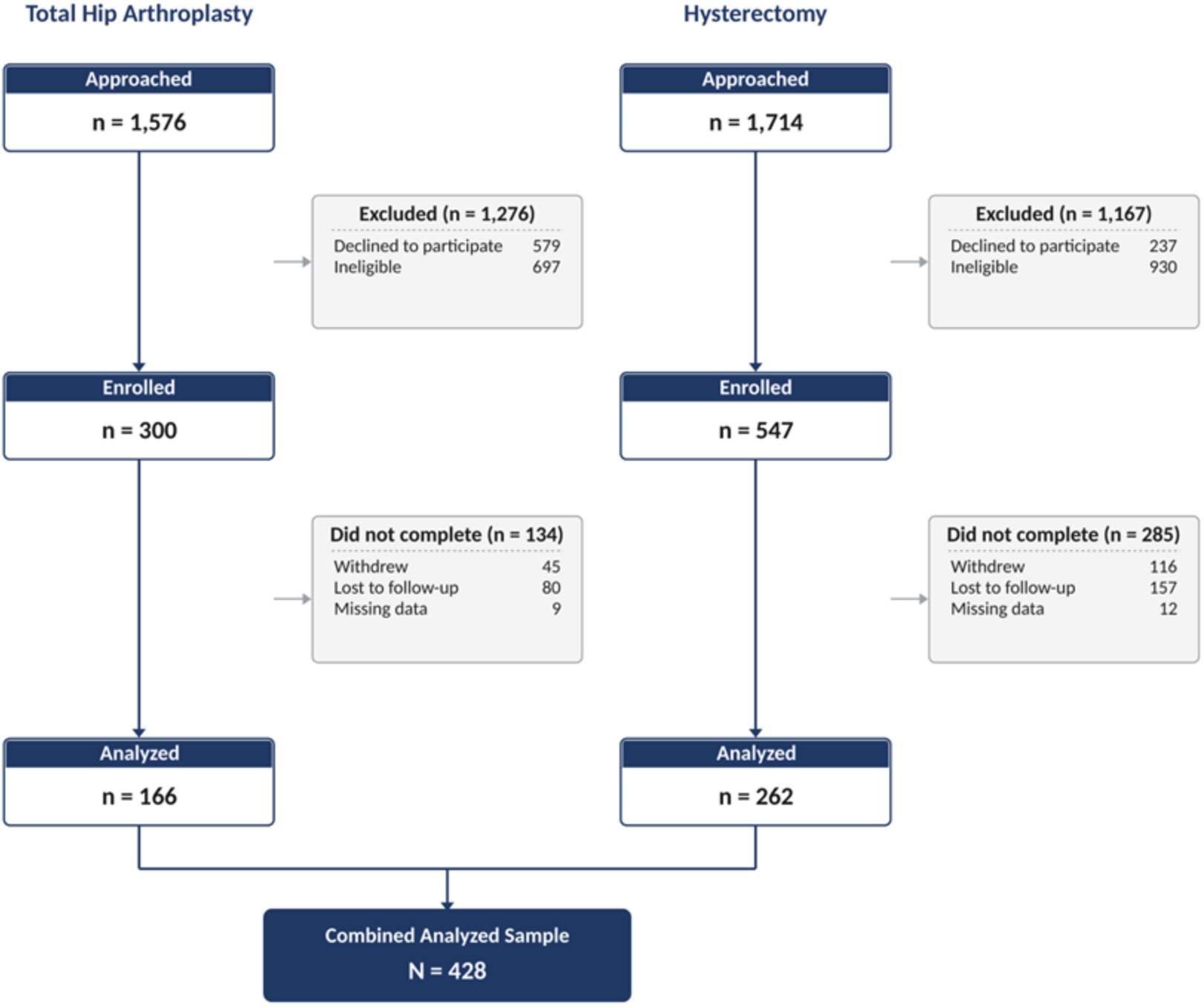
CONSORT flow diagram of participant enrollment, follow-up, and analysis.

**Figure 2.**
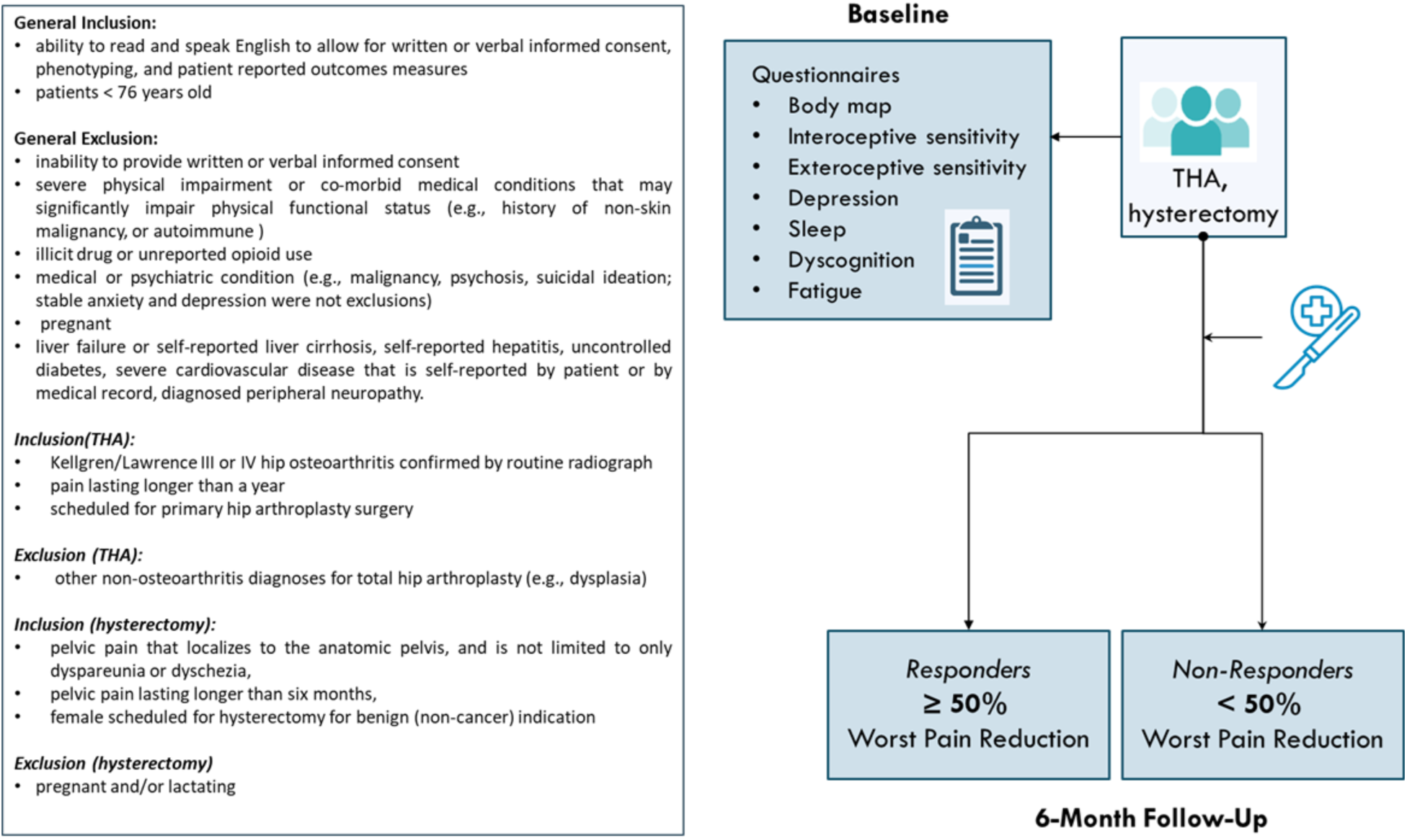
Inclusion and exclusion criteria for each cohort (left panel) and study design overview (right panel). Prior to surgery participants completed a battery of patient reported outcome measures and were assessed for a 50% reduction in worst pain six months after surgery. THA = Total Hip Arthroplasty.

### Primary Outcome: Surgical Analgesic Response

Study participants from both cohorts underwent surgical procedures as part of their standard clinical care. Pain levels were self-reported by participants using a 0-10 numeric rating scale, indicating their maximum (“worst”) pain intensity over the past four weeks both prior to and six months following treatment. For the purposes of these analyses, non-responsiveness to treatment was defined by a pain reduction of less than 50% at 6-months follow-up compared to baseline, following the Initiative on Methods, Measurement, and Pain Assessment in Clinical Trials (IMMPACT) guidelines for chronic pain outcomes.^12^ Participants were periodically contacted to complete outcomes measures (1 month, 3 months, 6 months) with the primary outcome designated at the six-month follow-up.

### Secondary Outcome: Patient Global Impression of Change

The Patient Global Impression of Change (PGIC) scale was administered concurrently with the worst pain outcome at 6-months follow-up. The PGIC asks patients to rate overall change in pain (“Since your surgery, your PAIN OVERALL is:”) from seven options: “very much improved,” “much improved,” “minimally improved,” “no change”, “minimally worse,” “much worse,” and “very much worse.” PGIC scales are frequently employed in clinical trials and are generally valued by patient partners as an alternative to numeric rating scales in addition to being a recommended component by IMMPACT guidelines.^13^

### Patient-Reported Outcome Measures

Symptoms were measured using item sets from validated patient-reported outcome measures, including the Identify Chronic Migraine Screener (ID-CM),^14^ Complex Medical Symptoms Inventory (CMSI),^15^ and Brief Self-Report Measure on Cognitive Dysfunction in Fibromyalgia.^16^ Additional assessments related to fatigue (FM),^17^ depression (8a),^18, 19^ and sleep related impairment (8a)^20^ were included from the PROMIS item banks. In total, 68 items were included in these analyses. The body map used was derived from the Michigan Body Map, which was further reduced into seven anatomically distinct regions, following our previous work examining the Generalized Sensory Sensitivity construct.^21–24^

A full list of these items can be found in **Supplemental Table 1**. These items can be mapped to seven larger symptom domains: sleep problems, negative affect, cognitive dysfunction, low energy/fatigue, widespread pain, and heightened interoceptive and exteroceptive sensitivity, each of which have been associated with chronic pain, chronic overlapping conditions (COPCs), development of new-onset pain, and poor treatment outcomes.^5–7, 21, 25-^27

At the baseline visit, prior to any surgical intervention, participants completed the 2011/2016 Fibromyalgia Survey Criteria^9, 10^ the Central Sensitization Inventory (CSI)^28^ and the PainDETECT.^11^ The Fibromyalgia Survey Criteria include a widespread pain index and a symptom severity scale that when combined can be used to calculate an overall score ranging from 0-31. The Central Sensitization Inventory assesses 25 health-related symptoms, with total overall scores ranging from 0 −100. PainDETECT is an assessment for the presence of neuropathic pain characteristics with scores ranging from −1 to 38 where a score of 12 or below represents a negative indication for neuropathic pain, scores between 13 and 18 are possible neuropathic pain, and a score of 19 or greater is probable neuropathic pain. The PainDETECT, despite not being a putative measure of nociplastic pain, has been shown to be associated with features of central sensitization in osteoarthritis and to predict surgical outcomes in this condition.^29, 30^

### Statistical Analyses

### Elastic Net Regression and Measure Construction

An elastic net regression with 10-fold cross-validation was utilized to help identify items that were most strongly associated with treatment non-responsiveness. To ensure that a range of items were included in the model output while limiting less significant items, an alpha regularization term of 0.5 was selected. This cross-validation technique allowed for the selection of a penalty parameter, lambda, that minimizes prediction error. This alpha regularization value was selected to maximize predictability while keeping the instrument concise and removing items with minimal impact. We also examined the median number of variables included in models as the alpha regularization value increased, and their relationship with the percentage of deviance explained, to help guide the final number of items retained. We also undertook a sensitivity analysis limiting inclusion to those who reported a baseline pain value of 3 or more. Final item selection was guided by comparing results from the primary and sensitivity analyses to retain items that were highly predictive in both scenarios.

### Measure Comparison

Comparisons of our derived measure (GAUGE) with established measures (i.e., Fibromyalgia Survey Criteria, Central Sensitization Inventory, and PainDETECT) utilized Akaike Information Criterion (AIC), Bayesian Information Criterion (BIC), area under the curve (AUC) analyses, and risk ratios for failure to achieve a 50% reduction in “worst pain” improvement over time. For analyses of PGIC values, ordinal regression was used with AIC and BIC values for model performance comparisons. Statistical analyses were performed in R (v4.1.1).

### Validation Sample

Following construction of the GAUGE screener, we tested its performance against the FM survey criteria in a sample of total knee arthroplasty patients recruited through the Michigan Surgical Quality Collaborative clinical registry database (n=54). The primary outcome for this study is a 50% improvement in pain severity on the Brief Pain Inventory, a measure widely used to track analgesic response in clinical trials. AIC, BIC, and AUC were compared between GAUGE summary scores and the FM Survey Criteria. The PainDETECT and Central Sensitization Inventory were not available in this study.

## RESULTS

### Demographics

A total of 428 participants were included (166 THA and 262 hysterectomy), with a mean age of 50.97 years; 80.61% were female and 35% failed to achieve a 50% pain reduction (**Table 1**).

**Table 1.** Participant demographics.

|  | All Participants | Responders | Non-Responders | Total Hip Arthroplasty | Hysterectomy |
| --- | --- | --- | --- | --- | --- |
| **N** | 428 | 277 | 151 | 166 | 262 |
| **Age (years)** | 50.97 (12.70) | 51.80 (12.90) | 49.40 (12.20) | 62.90 (8.57) | 43.40 (8.28) |
| **Male** | 83 (19.39%) | 58 (20.94%) | 25 (16.56%) | 83 (50.00%) | N/A |
| **Female** | 345 (80.61%) | 219 (79.06%) | 126 (83.44%) | 83 (50.00%) | 262 (100.00%) |
| **7-Region Body Map** | 2.19 (1.76) | 1.82 (1.50) | 2.86 (1.98) | 2.58 (1.31) | 1.94 (1.95) |
| **Fibromyalgia Survey Criteria** | 7.96 (4.88) | 6.91 (4.02) | 9.89 (5.68) | 6.50 (3.38) | 8.88 (5.43) |
| **Central Sensitization Inventory** | 34.28 (16.66) | 30.90 (15.20) | 40.50 (17.40) | 25.50 (12.40) | 39.80 (16.70) |
| **PainDETECT** | 7.71 (6.22) | 6.83 (6.00) | 9.34 (6.30) | 8.11 (5.80) | 7.47 (6.47) |

**Note:** Values are mean (SD) unless otherwise indicated. Participant sex is reported as n (%). Responders are defined as participants with a 50% or greater decrease in their strongest pain between baseline and follow-up visit.

### Model Fit by Item Count

Gains in percent deviance explained plateaued after the first five items, with only marginal additional improvement once 12 or more items were included (**Supplemental Figure 1**).

### Item Identification

Elastic net regression identified 11 items from the 68 candidate items that were associated with analgesic non-response and retained in both primary and sensitivity analyses. Seven corresponded to regions on a body map, each scored for the presence or absence of pain, and four were binary (yes/no) symptom items from the CMSI: two related to interoception (“numbness or tingling in arms or legs” and “nausea“) and two related to exteroception (“sensitivity to sound” and “sensitivity to odors“). The GAUGE summary score sums these items for a possible range of 0–11. Full elastic net results are presented in **Supplemental Table 2**. The final GAUGE screener items appear in **Supplemental Figure 2**.

### Association with Existing Measures

The GAUGE screener was most strongly associated with the Fibromyalgia Survey Criteria (r = 0.75), followed by the Central Sensitization Inventory (r = 0.52) and PainDETECT (r = 0.52).

### Analysis of Constituent Subdomains

GAUGE items represent three correlated but distinct subdomains: the spatial distribution of pain on a body map, interoceptive sensitivity, and exteroceptive sensitivity. We examined whether each subdomain provided unique predictive value relative to the full summary measure. Models were fit using each subdomain alone (e.g., body map only), pairwise combinations (e.g., interoception items + body map), and the full summary measure combining all three. The full summary measure outperformed every single-subdomain and pairwise model across all model fit metrics (**Supplemental Table 3**).

### Model Performance

The GAUGE summary score was strongly associated with the proportion of participants achieving a 50% reduction in pain following surgery (R² = 0.87; **Figure 3**). In comparative model fit, GAUGE outperformed the Fibromyalgia Survey Criteria, Central Sensitization Inventory, and PainDETECT for predicting surgical non-response, yielding the lowest AIC (496.02) and BIC (504.14), the highest AUC (0.7384), and the largest risk ratio (1.5350; 95% CI 1.3421–1.7555; **Table 2**). GAUGE achieved a sensitivity of 0.741 and a specificity of 0.635 for identifying surgical non-responders. The advantage over the next-best comparator was substantial (ΔAIC = ΔBIC = 26.95).

**Figure 3.**
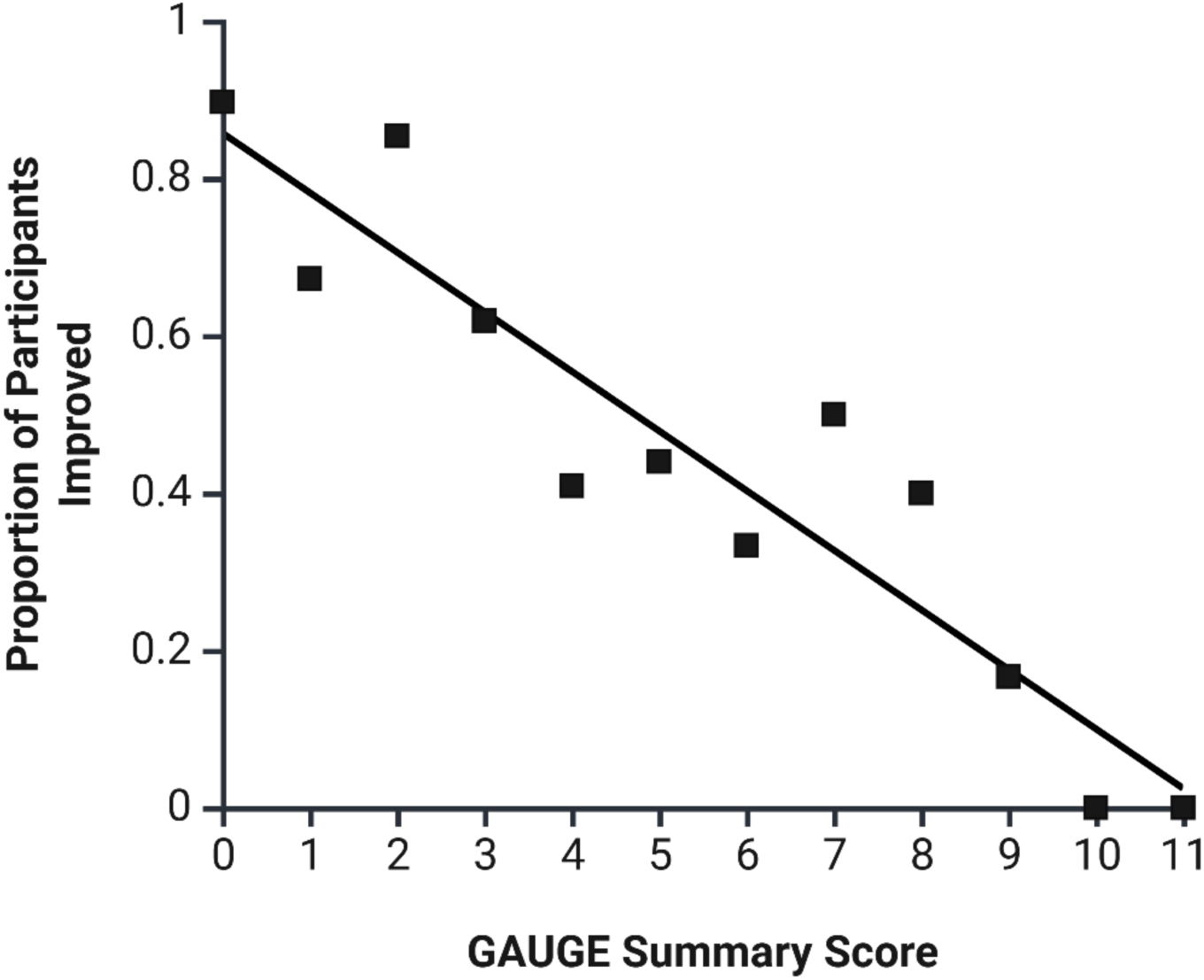
Scatterplot showing the association between GAUGE scores (0-11) and the proportion participants achieving a 50% reduction in worst pain (e.g., 90% of participants scoring a one [1] on the GAUGE were responders.

**Table 2.** Model performance for predicting surgical analgesic non-response.

|  | AIC | BIC | AUC | Risk Ratio (95% CI) |
| --- | --- | --- | --- | --- |
| **GAUGE Summary Measure** | **496.02** | **504.14** | **0.7384** | **1.5350 (1.3421, 1.7555)** |
| Fibromyalgia Survey Criteria | 522.97 | 531.09 | 0.6613 | 1.3699 (1.2062, 1.5559) |
| Central Sensitization Inventory | 526.14 | 534.26 | 0.6598 | 1.4218 (1.2241, 1.6516) |
| PainDETECT | 543.98 | 552.10 | 0.6369 | 1.2526 (1.0945, 1.4564) |

**Note:** AIC = Akaike information criterion; BIC = Bayesian information criterion; AUC = area under the receiver operating characteristic curve; CI = confidence interval. Lower AIC and BIC and higher AUC indicate better model performance. The GAUGE summary measure (highlighted) outperformed each individual instrument across all criteria.

The same four measures were compared as predictors of PGIC (**Table 3**). GAUGE again yielded the lowest AIC (934.87) and BIC (963.28). Its advantage over the Fibromyalgia Survey Criteria however was small (ΔAIC = 1.52; ΔBIC = 1.62), whereas GAUGE clearly outperformed the Central Sensitization Inventory (ΔAIC = 10.94; ΔBIC = 10.95) and PainDETECT (ΔAIC = 18.32; ΔBIC = 18.32).

**Table 3.** Model performance for predicting Patient Global Impression of Change following surgery.

| Measure | AIC | BIC |
| --- | --- | --- |
| **GAUGE Summary Measure** | **934.87** | **963.28** |
| Fibromyalgia Survey Criteria | 936.39 | 964.90 |
| Central Sensitization Inventory | 945.81 | 974.23 |
| PainDETECT | 953.19 | 981.60 |

**Note:** AIC = Akaike information criterion; BIC = Bayesian information criterion. Lower values indicate better model performance.

### Model Validation

In an independent validation cohort of 54 total knee arthroplasty patients (**Table 4**), GAUGE showed numerically better values than the Fibromyalgia Survey Criteria on all three metrics (AIC = 69.90 vs. 70.95; BIC = 73.88 vs. 74.93; AUC = 0.6760 vs. 0.6566), although the differences were small (ΔAIC = 1.05; ΔBIC = 1.05; ΔAUC = 0.0194) and both measures showed only modest discrimination.

**Table 4.** Model performance for predicting pain severity on the Brief Pain Inventory in a total knee arthroplasty validation cohort (n = 54).

| Measure | AIC | BIC | AUC |
| --- | --- | --- | --- |
| **GAUGE Summary Measure** | **69.90** | **73.88** | **0.6760** |
| Fibromyalgia Survey Criteria | 70.95 | 74.93 | 0.6566 |

**Note:** AIC = Akaike information criterion; BIC = Bayesian information criterion; AUC = area under the receiver operating characteristic curve; Lower AIC and BIC and higher AUC indicate better model performance.

## DISCUSSION

Leveraging data from two large cohorts of well-phenotyped surgical patients, we used a data-driven item-selection approach to develop a brief screener for poor analgesic response to surgery. The resulting GAUGE screener captures three symptom subdomains — spatial distribution of pain, interoceptive sensitivity (internal bodily sensations), and exteroceptive sensitivity (environmental stimuli) — that together constitute the previously described Generalized Sensory Sensitivity construct.^21, 25^ Patients with higher GAUGE scores, indicating widespread pain and heightened multisensory sensitivity, were substantially less likely to achieve 50% pain relief following surgery and scores were also predictive of patient-reported improvement on the PGIC. Additionally, GAUGE outperformed the Fibromyalgia Survey Criteria, Central Sensitization Inventory, and PainDETECT for predicting surgical non-response, and these findings were robust to sensitivity analyses. In a small total knee arthroplasty validation cohort, GAUGE showed good performance compared to the Fibromyalgia Survey Criteria. GAUGE offers several advantages over available tools: items were selected through a data-driven procedure, the resulting subdomains have a clear theoretical and neurobiological basis, and the measure is brief enough for routine pre-surgical use.

Nociplastic pain is an emerging construct related to CNS sensitization, referring to pain that cannot be readily attributed to peripheral tissue damage, inflammation, or neuropathic injury.^31^ The International Association for the Study of Pain has identified nociplastic pain as a priority area, because when nociplastic mechanisms contribute substantially to a patient’s pain, therapies targeting peripheral pain generators, including surgery, are less likely to succeed. The current findings demonstrate that this risk can be efficiently captured through a self-report screener of nociplastic pain symptoms.

Previous studies examining self-report measures of central sensitization or nociplastic pain symptoms and surgical outcomes have been limited because available measures were not designed for screening surgical populations. Although the current analyses considered seven candidate symptom domains, the three retained subdomains — pain distribution, internal bodily sensitivity, and external environmental sensitivity — correspond directly to the constituents of the Generalized Sensory Sensitivity construct. Assessing all three subdomains produced more accurate risk profiles than any single subdomain alone, consistent with the hypothesis that these symptoms reflect a shared underlying mechanism rather than independent processes. Each of these symptoms are strongly associated with the neurobiological sensitization believed to undergird multiple clinically defined pain conditions.^26, 27, 32^ Altered patterns of activation and functional connectivity involving the insula, a key sensory integration brain structure, have been consistently noted in neuroimaging studies examining these symptoms.^33–36^ These neurobiological findings, coupled with the current results, support a theoretical model in which entrenched nociplastic pain mechanisms maintain a chronic pain state even when peripheral pain drivers have been surgically corrected or removed.

Several limitations should be noted. First, although we examined two surgical cohorts with different etiologies for pain, the full range of surgical procedures undertaken for pain relief may not show the relationships observed here; validation in additional surgical populations will be necessary. Second, the external validation cohort was small (n=54), and the magnitude of GAUGE’s advantage over the Fibromyalgia Survey Criteria was modest in that sample. Replication in larger validation cohorts is needed before clinical adoption. Third, our development sample was predominantly female (80.6%) and drawn from two specific surgical indications, which may limit generalizability. Finally, the focus on a 50% improvement in worst pain and PGIC, while consistent with frameworks for interpreting clinical trials, may not capture relationships between GAUGE and other relevant outcomes such as functional recovery or opioid use.

## Conclusions

GAUGE is a brief, theoretically grounded, and data-driven screener associated with poor analgesic response to surgery. It has potential utility for enriching clinical trials of analgesic interventions, providing personalized pre-surgical risk assessment, and informing planning for adjuvant pain-relieving treatments in patients identified with elevated risk.

## Disclosures

This project was supported by funding from NIH (P50AR070600 to DJC and CB and R01HD088712 to SA and DJC).

## Data Availability

All data produced in the present study are available upon reasonable request to the authors.

## Notes

### Competing Interest Statement

The authors have declared no competing interest.

### Author Declarations

IRB of University of Michigan gave ethical approval for this work.

## REFERENCES

1. Wright JD, Huang Y, Li AH, Melamed A, Hershman DL. Nationwide Estimates of Annual Inpatient and Outpatient Hysterectomies Performed in the United States. Obstetrics & Gynecology. 2022;139(3):446–8. doi: 10.1097/aog.0000000000004679. PubMed PMID: 00006250-202203000-00014.

2. Cram P, Lu X, Kates SL, Singh JA, Li Y, Wolf BR. Total Knee Arthroplasty Volume, Utilization, and Outcomes Among Medicare Beneficiaries, 1991-2010. JAMA. 2012;308(12):1227–36. doi: 10.1001/2012.jama.11153.

3. Brandsborg B, Dueholm M, Nikolajsen L, Kehlet H, Jensen TS. A prospective study of risk factors for pain persisting 4 months after hysterectomy. Clin J Pain. 2009;25(4):263–8. Epub 2009/07/11. doi: 10.1097/AJP.0b013e31819655ca. PubMed PMID: 19590472.

4. Beswick AD, Wylde V, Gooberman-Hill R, Blom A, Dieppe P. What proportion of patients report long-term pain after total hip or knee replacement for osteoarthritis? A systematic review of prospective studies in unselected patients. BMJ Open. 2012;2(1):e000435. Epub 2012/02/24. doi: 10.1136/bmjopen-2011-000435. PubMed PMID: 22357571; PMCID: PMC3289991 any of the authors that could create a potential conflict of interest with regard to the work.

5. Brummett CM, Urquhart AG, Hassett AL, Tsodikov A, Hallstrom BR, Wood NI, Williams DA, Clauw DJ. Characteristics of Fibromyalgia Independently Predict Poorer Long-Term Analgesic Outcomes Following Total Knee and Hip Arthroplasty. Arthritis & Rheumatology. 2015;67(5):1386–94. doi: 10.1002/art.39051.

6. As-Sanie S, Till SR, Schrepf AD, Griffith KC, Tsodikov A, Missmer SA, Clauw DJ, Brummett CM. Incidence and predictors of persistent pelvic pain following hysterectomy in women with chronic pelvic pain. American journal of obstetrics and gynecology. 2021;225(5):568. e1–. e11.

7. Orr NL, Huang AJ, Liu YD, Noga H, Bedaiwy MA, Williams C, Allaire C, Yong PJ. Association of central sensitization inventory scores with pain outcomes after endometriosis surgery. JAMA Network Open. 2023;6(2):e230780–e.

8. Mayer TG, Neblett R, Cohen H, Howard KJ, Choi YH, Williams MJ, Perez Y, Gatchel RJ. The development and psychometric validation of the central sensitization inventory. Pain Practice. 2012;12(4):276–85.

9. Wolfe F, Clauw DJ, Fitzcharles M-A, Goldenberg DL, Häuser W, Katz RL, Mease PJ, Russell AS, Russell IJ, Walitt B, editors. 2016 Revisions to the 2010/2011 fibromyalgia diagnostic criteria. Seminars in arthritis and rheumatism; 2016: Elsevier.

10. Wolfe F, Clauw DJ, Fitzcharles MA, Goldenberg DL, Hauser W, Katz RS, Mease P, Russell AS, Russell IJ, Winfield JB. Fibromyalgia criteria and severity scales for clinical and epidemiological studies: a modification of the ACR Preliminary Diagnostic Criteria for Fibromyalgia. J Rheumatol. 2011;38(6):1113–22. Epub 2011/02/03. doi: 10.3899/jrheum.100594. PubMed PMID: 21285161.

11. Freynhagen R, Baron R, Gockel U, Tolle TR. painDETECT: a new screening questionnaire to identify neuropathic components in patients with back pain. Curr Med Res Opin. 2006;22(10):1911–20. Epub 2006/10/07. doi: 10.1185/030079906X132488. PubMed PMID: 17022849.

12. Smith SM, Dworkin RH, Turk DC, McDermott MP, Eccleston C, Farrar JT, Rowbotham MC, Bhagwagar Z, Burke LB, Cowan P, Ellenberg SS, Evans SR, Freeman RL, Garrison LP, Iyengar S, Jadad A, Jensen MP, Junor R, Kamp C, Katz NP, Kesslak JP, Kopecky EA, Lissin D, Markman JD, Mease PJ, O’Connor AB, Patel KV, Raja SN, Sampaio C, Schoenfeld D, Singh J, Steigerwald I, Strand V, Tive LA, Tobias J, Wasan AD, Wilson HD. Interpretation of chronic pain clinical trial outcomes: IMMPACT recommended considerations. Pain. 2020;161(11):2446–61. Epub 2020/06/11. doi: 10.1097/j.pain.0000000000001952. PubMed PMID: 32520773; PMCID: PMC7572524.

13. Dworkin RH, Turk DC, Farrar JT, Haythornthwaite JA, Jensen MP, Katz NP, Kerns RD, Stucki G, Allen RR, Bellamy N, Carr DB, Chandler J, Cowan P, Dionne R, Galer BS, Hertz S, Jadad AR, Kramer LD, Manning DC, Martin S, McCormick CG, McDermott MP, McGrath P, Quessy S, Rappaport BA, Robbins W, Robinson JP, Rothman M, Royal MA, Simon L, Stauffer JW, Stein W, Tollett J, Wernicke J, Witter J. Core outcome measures for chronic pain clinical trials: IMMPACT recommendations. Pain. 2005;113(1-2):9–19. Epub 2004/12/29. doi: 10.1016/j.pain.2004.09.012. PubMed PMID: 15621359.

14. Pavlovic JM, Yu JS, Silberstein SD, Reed ML, Cowan RP, Dabbous F, Pulicharam R, Viswanathan HN, Lipton RB. Evaluation of the 6-item Identify Chronic Migraine screener in a large medical group. Headache. 2021;61(2):335–42. Epub 2021/01/10. doi: 10.1111/head.14035. PubMed PMID: 33421098; PMCID: PMC7986415.

15. Williams DA, Schilling S. Advances in the assessment of fibromyalgia. Rheum Dis Clin North Am. 2009;35(2):339–57. Epub 2009/08/04. doi: 10.1016/j.rdc.2009.05.007. PubMed PMID: 19647147; PMCID: PMC2721827.

16. Kratz AL, Schilling SG, Goesling J, Williams DA. Development and initial validation of a brief self-report measure of cognitive dysfunction in fibromyalgia. J Pain. 2015;16(6):527–36. Epub 2015/03/10. doi: 10.1016/j.jpain.2015.02.008. PubMed PMID: 25746197; PMCID: PMC4456217.

17. Kratz AL, Schilling S, Goesling J, Williams DA. The PROMIS FatigueFM Profile: a self-report measure of fatigue for use in fibromyalgia. Qual Life Res. 2016;25(7):1803–13. Epub 2016/01/30. doi: 10.1007/s11136-016-1230-9. PubMed PMID: 26821919; PMCID: PMC4893882.

1818 Cella D, Riley W, Stone A, Rothrock N, Reeve B, Yount S, Amtmann D, Bode R, Buysse D, Choi S, Cook K, Devellis R, DeWalt D, Fries JF, Gershon R, Hahn EA, Lai JS, Pilkonis P, Revicki D, Rose M, Weinfurt K, Hays R, Group PC. The Patient-Reported Outcomes Measurement Information System (PROMIS) developed and tested its first wave of adult self-reported health outcome item banks: 2005-2008. J Clin Epidemiol. 2010;63(11):1179–94. Epub 2010/08/06. doi: 10.1016/j.jclinepi.2010.04.011. PubMed PMID: 20685078; PMCID: PMC2965562.

1919 Pilkonis PA, Choi SW, Reise SP, Stover AM, Riley WT, Cella D, Group PC. Item banks for measuring emotional distress from the Patient-Reported Outcomes Measurement Information System (PROMIS®): depression, anxiety, and anger. Assessment. 2011;18(3):263–83.

20. Yu L, Buysse DJ, Germain A, Moul DE, Stover A, Dodds NE, Johnston KL, Pilkonis PA. Development of short forms from the PROMIS™ sleep disturbance and sleep-related impairment item banks. Behavioral sleep medicine. 2012;10(1):6–24.

21. Schrepf A, Hellman KM, Bohnert AM, Williams DA, Tu FF. Generalized sensory sensitivity is associated with comorbid pain symptoms: a replication study in women with dysmenorrhea. Pain. 2023;164(1):142–8.

22. Adams MC, Brummett CM, Wandner LD, Topaloglu U, Hurley RW. Michigan body map: connecting the NIH HEAL IMPOWR network to the HEAL ecosystem. Pain Medicine. 2023;24(7):907–9.

23. Brummett CM, Bakshi RR, Goesling J, Leung D, Moser SE, Zollars JW, Williams DA, Clauw DJ, Hassett AL. Preliminary validation of the Michigan body map. Pain. 2016;157(6):1205–12.

24. Hassett AL, Pierce J, Goesling J, Fritsch L, Bakshi RR, Kohns DJ, Brummett CM. Initial validation of the electronic form of the Michigan Body Map. Regional Anesthesia & Pain Medicine. 2020;45(2):145–50.

25. Schrepf A, Williams DA, Gallop R, Naliboff BD, Basu N, Kaplan C, Harper DE, Landis JR, Clemens JQ, Strachan E. Sensory sensitivity and symptom severity represent unique dimensions of chronic pain: a MAPP Research Network study. Pain. 2018;159(10):2002–11.

26. Bair E, Ohrbach R, Fillingim RB, Greenspan JD, Dubner R, Diatchenko L, Helgeson E, Knott C, Maixner W, Slade GD. Multivariable Modeling of Phenotypic Risk Factors for First-Onset TMD: The OPPERA Prospective Cohort Study. The Journal of Pain. 2013;14(12, Supplement):T102–T15. doi: 10.1016/j.jpain.2013.09.003.

27. Ohrbach R, Sharma S, Fillingim RB, Greenspan JD, Rosen JD, Slade GD. Clinical Characteristics of Pain Among Five Chronic Overlapping Pain Conditions. J Oral Facial Pain Headache. 2020;34(Suppl):s29–s42. Epub 2020/09/26. doi: 10.11607/ofph.2573. PubMed PMID: 32975539; PMCID: PMC10073942.

28. Neblett R, Cohen H, Choi Y, Hartzell MM, Williams M, Mayer TG, Gatchel RJ. The Central Sensitization Inventory (CSI): establishing clinically significant values for identifying central sensitivity syndromes in an outpatient chronic pain sample. J Pain. 2013;14(5):438–45. Epub 2013/03/16. doi: 10.1016/j.jpain.2012.11.012. PubMed PMID: 23490634; PMCID: PMC3644381.

29. Soni A, Wanigasekera V, Mezue M, Cooper C, Javaid MK, Price Andrew J, Tracey I. Central Sensitization in Knee Osteoarthritis: Relating Presurgical Brainstem Neuroimaging and PainDETECT-Based Patient Stratification to Arthroplasty Outcome. Arthritis & Rheumatology. 2019;71(4):550–60. doi: 10.1002/art.40749.

30. Kurien T, Arendt-Nielsen L, Petersen KK, Graven-Nielsen T, Scammell BE. Preoperative Neuropathic Pain-like Symptoms and Central Pain Mechanisms in Knee Osteoarthritis Predicts Poor Outcome 6 Months After Total Knee Replacement Surgery. The Journal of Pain. 2018;19(11):1329–41. doi: 10.1016/j.jpain.2018.05.011.

31. Kaplan CM, Kelleher E, Irani A, Schrepf A, Clauw DJ, Harte SE. Deciphering nociplastic pain: clinical features, risk factors and potential mechanisms. Nat Rev Neurol. 2024;20(6):347–63. Epub 2024/05/17. doi: 10.1038/s41582-024-00966-8. PubMed PMID: 38755449.

32. Schrepf A, Williams DA, Gallop R, Naliboff BD, Basu N, Kaplan C, Harper DE, Landis JR, Clemens JQ, Strachan E, Griffith JW, Afari N, Hassett A, Pontari MA, Clauw DJ, Harte SE. Sensory sensitivity and symptom severity represent unique dimensions of chronic pain: a MAPP Research Network study. Pain. 2018;159(10):2002–11. Epub 2018/06/05. doi: 10.1097/j.pain.0000000000001299. PubMed PMID: 29863527; PMCID: PMC6705610.

33. Boeckle M, Schrimpf M, Liegl G, Pieh C. Neural correlates of somatoform disorders from a meta-analytic perspective on neuroimaging studies. NeuroImage: Clinical. 2016;11:606–13. doi: 10.1016/j.nicl.2016.04.001.

34. López-Solà M, Pujol J, Wager TD, Garcia-Fontanals A, Blanco-Hinojo L, Garcia-Blanco S, Poca-Dias V, Harrison BJ, Contreras-Rodríguez O, Monfort J, Garcia-Fructuoso F, Deus J. Altered Functional Magnetic Resonance Imaging Responses to Nonpainful Sensory Stimulation in Fibromyalgia Patients. Arthritis & Rheumatology. 2014;66(11):3200–9. doi: 10.1002/art.38781.

35. van Ettinger-Veenstra H, Lundberg P, Alföldi P, Södermark M, Graven-Nielsen T, Sjörs A, Engström M, Gerdle B. Chronic widespread pain patients show disrupted cortical connectivity in default mode and salience networks, modulated by pain sensitivity. J Pain Res. 2019;12:1743–55. Epub 2019/06/20. doi: 10.2147/jpr.S189443. PubMed PMID: 31213886; PMCID: PMC6549756.

36. Ichesco E, Schmidt-Wilcke T, Bhavsar R, Clauw DJ, Peltier SJ, Kim J, Napadow V, Hampson JP, Kairys AE, Williams DA, Harris RE. Altered Resting State Connectivity of the Insular Cortex in Individuals With Fibromyalgia. The Journal of Pain. 2014;15(8):815–26.e1. doi: 10.1016/j.jpain.2014.04.007.

