## Supplemental Materials for "Generalized Sensory Sensitivity for Prediction of Post-Surgical Analgesic Outcomes: An Observational Cohort Study of Total Hip Arthroplasty and Hysterectomy"

**Supplemental Table 1:** Items and measures considered for construct development. Items shaded blue are those retained in the final model and form the GAUGE screener.

| Item | Instrument |
|---|---|
| Participant age at baseline | - |
| Participant sex at birth | - |
| **7-Region Body map** | |
| Strongest pain at baseline | |
| In the last 3 months (past 90 days), on how many days did you have a headache of any type? | Identify Chronic Migraine Screener |
| In the last month (past 30 days), on how many days did you have a headache of any type? | Identify Chronic Migraine Screener |
| How often were you unusually sensitive to light (e.g., you felt more comfortable in a dark place)? | Identify Chronic Migraine Screener |
| How often were you unusually sensitive to sound (e.g., you felt more comfortable in a quiet place)? | Identify Chronic Migraine Screener |
| How often was the pain moderate or severe? | Identify Chronic Migraine Screener |
| How often did you feel nauseated or sick to your stomach? | Identify Chronic Migraine Screener |
| Muscle spasms 3 months last year | Complex Medical Symptoms Inventory |
| Recurrent fevers 3 months last year | Complex Medical Symptoms Inventory |
| Dry eyes 3 months last year | Complex Medical Symptoms Inventory |
| Dry mouth 3 months last year | Complex Medical Symptoms Inventory |
| Fingers turn blue and/or white in the cold 3 months last year | Complex Medical Symptoms Inventory |
| **Numbness or tingling in arms or legs 3 months last year** | **Complex Medical Symptoms Inventory** |
| Shortness of breath during normal activity 3 months last year | Complex Medical Symptoms Inventory |
| Palpitations 3 months last year | Complex Medical Symptoms Inventory |
| Rapid heart rate 3 months last year | Complex Medical Symptoms Inventory |
| Heartburn 3 months last year | Complex Medical Symptoms Inventory |
| Vomiting 3 months last year | Complex Medical Symptoms Inventory |
| **Nausea 3 months last year** | **Complex Medical Symptoms Inventory** |
| Problems with balance 3 months last year | Complex Medical Symptoms Inventory |
| Dizziness 3 months last year | Complex Medical Symptoms Inventory |
| Sensation of ear blockage or fullness 3 months last year | Complex Medical Symptoms Inventory |
| Sinus pressure 3 months last year | Complex Medical Symptoms Inventory |
| Sensitivity to certain chemical, such as perfumes, laundry detergents, gasoline and others 3 months last year | Complex Medical Symptoms Inventory |
| **Sensitivity to sound 3 months last year** | **Complex Medical Symptoms Inventory** |
| **Sensitivity to odors 3 months last year** | **Complex Medical Symptoms Inventory** |
| Frequent sensitivity to bright lights 3 months last year | Complex Medical Symptoms Inventory |
| How tired did you feel on average? | PROMIS Fatigue |
| How fatigued were you on average? | PROMIS Fatigue |
| How exhausted were you on average? | PROMIS Fatigue |
| To what degree did fatigue interfere with your social activities? | PROMIS Fatigue |
| To what degree did fatigue interfere with your recreational activities? | PROMIS Fatigue |
| To what degree did you have trouble starting things because of fatigue? | PROMIS Fatigue |
| To what degree did you have trouble finishing things because of fatigue? | PROMIS Fatigue |
| To what degree did fatigue make it difficult to make decisions? | PROMIS Fatigue |
| To what degree did fatigue make you feel slowed down in your thinking? | PROMIS Fatigue |

| | |
|---|---|
| How often were you less effective at home due to fatigue? | PROMIS Fatigue |
| How often did you have to push yourself to get things done because of your fatigue? | PROMIS Fatigue |
| How often did you have to limit your social activities because of fatigue? | PROMIS Fatigue |
| How often were you too tired to socialize with your friends? | PROMIS Fatigue |
| How often were you too tired to think clearly? | PROMIS Fatigue |
| How often did fatigue make you more forgetful? | PROMIS Fatigue |
| What was the level of your fatigue on most days? | PROMIS Fatigue |
| I have been able to think clearly without extra effort. | Brief Self-Report Measure on Cognitive Dysfunction in Fibromyalgia |
| My mind has been as sharp as usual. | Brief Self-Report Measure on Cognitive Dysfunction in Fibromyalgia |
| I have been able to remember things as easily as usual without extra effort. | Brief Self-Report Measure on Cognitive Dysfunction in Fibromyalgia |
| I have been able to learn new things easily, like telephone numbers or instructions. | Brief Self-Report Measure on Cognitive Dysfunction in Fibromyalgia |
| My ability to concentrate has been good. | Brief Self-Report Measure on Cognitive Dysfunction in Fibromyalgia |
| I have been able to pay attention and keep track of what I was doing without extra effort. | Brief Self-Report Measure on Cognitive Dysfunction in Fibromyalgia |
| I have had trouble shifting back and forth between different activities that require thinking. | Brief Self-Report Measure on Cognitive Dysfunction in Fibromyalgia |
| I had trouble planning out the steps of a task. | Brief Self-Report Measure on Cognitive Dysfunction in Fibromyalgia |
| I have had to work harder than usual to express myself clearly. | Brief Self-Report Measure on Cognitive Dysfunction in Fibromyalgia |
| I have had trouble finding the right word(s) to express myself. | Brief Self-Report Measure on Cognitive Dysfunction in Fibromyalgia |
| In the past 7 days…I felt depressed | PROMIS Depression |
| In the past 7 days…I felt nothing could cheer me up | PROMIS Depression |
| In the past 7 days…I felt worthless | PROMIS Depression |
| In the past 7 days…I felt unhappy | PROMIS Depression |
| I had a hard time getting things done because I was sleepy | PROMIS Sleep |
| I felt alert when I woke up | PROMIS Sleep |
| I felt tired | PROMIS Sleep |
| I had problems during the day because of poor sleep | PROMIS Sleep |
| I had a hard time concentrating because of poor sleep | PROMIS Sleep |
| I felt irritable because of poor sleep | PROMIS Sleep |
| I was sleepy during the daytime | PROMIS Sleep |
| I had trouble staying awake during the day | PROMIS Sleep |

**Note:** Items included in this analysis are from the following validated patient-reported outcome measures: PROMIS Sleep, Fatigue and Depression; Brief Self-Report Measure on Cognitive Dysfunction in Fibromyalgia; Identify Chronic Migraine Screener; and Complex Medical Symptoms Inventory.

**Supplemental Table 2:** Elastic net with K-fold cross validation results.

| Variable | Elastic Net – 50% Reduction in Baseline Pain | Elastic Net 50% Reduction in Baseline Pain (Worst Pain ≥ 3 at baseline) |
|---|---|---|
| Numbness or tingling in arms or legs (CMSI 10) | -0.1383 | -0.1134 |
| Sensitivity to sound (CMSI 36) | -0.1102 | -0.0722 |
| Number of painful regions | -0.0349 | -0.0408 |
| Nausea (CMSI 18) | -0.0342 | -0.0634 |
| Sensitivity to odors (CMSI 37) | -0.0167 | -0.0102 |
| Muscle spasms (CMSI 3) | -0.0066 | - |
| I felt depressed (DEP 29) | -0.0065 | - |
| ...on how many days did you have a headache of any type? (ID-CM 01) | -0.0009 | -0.0013 |
| Palpitations (CMSI 14) | - | -0.0181 |
| How often did you feel nauseated or sick to your stomach? (ID-CM 06) | - | -0.0038 |

**Note:** Values represent log odds. The presence or increase of each of the above variables are associated with greater odds of treatment non-responsiveness. A sensitivity analysis limited to participants with baseline pain values of 3 or more was included. Complex Medical Symptoms Inventory (CMSI); PROMIS Depression (DEP); Identify Chronic Migraine Screener (ID-CM)

**Supplemental Table 3.** Performance comparison of each GAUGE constituent subdomain and the full summary measure.

| **Subdomain(s)** | **AIC** | **BIC** | **AUC** |
|---|---|---|---|
| Body Map | 525.17 | 533.29 | 0.6821 |
| Interoception | 519.20 | 527.32 | 0.6381 |
| Exteroception | 528.60 | 536.72 | 0.5979 |
| Body Map + Interoception | 507.60 | 515.72 | 0.7201 |
| Body Map + Exteroception | 510.19 | 518.31 | 0.7134 |
| Interoception + Exteroception | 507.89 | 516.01 | 0.6617 |
| **GAUGE Summary Measure (full model)** | **496.02** | **504.14** | **0.7384** |

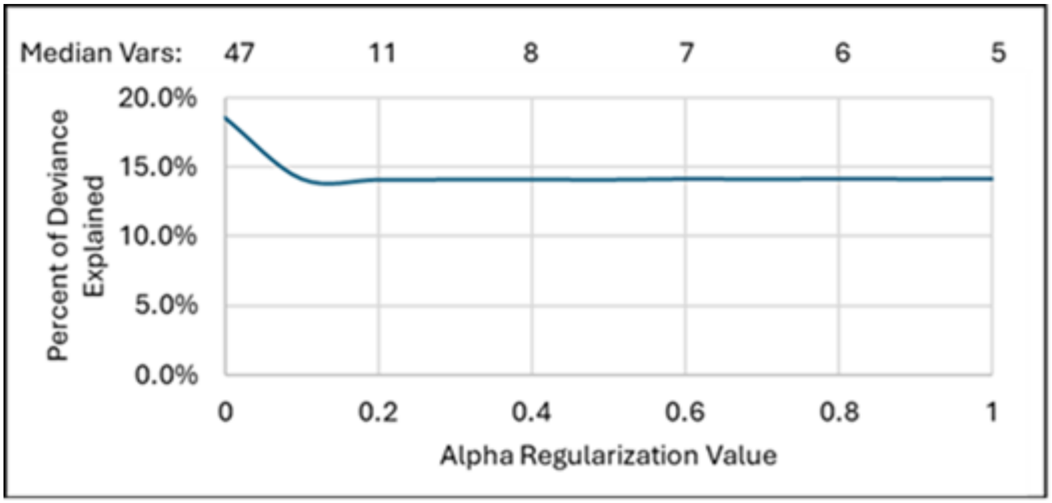


**Supplemental Figure 1.** Model parsimony versus fit across elastic net alpha values. Percent of deviance explained (y-axis) is plotted against the alpha regularization parameter (x-axis), with the median number of retained items shown above. Increasing alpha reduced model size from 47 to 5 items with minimal loss of explanatory power (~18% to ~14%).

**Supplemental Figure 2.** GAUGE screener.

The body map below is divided into seven regions. Please check each region where you have experienced pain during the last week.

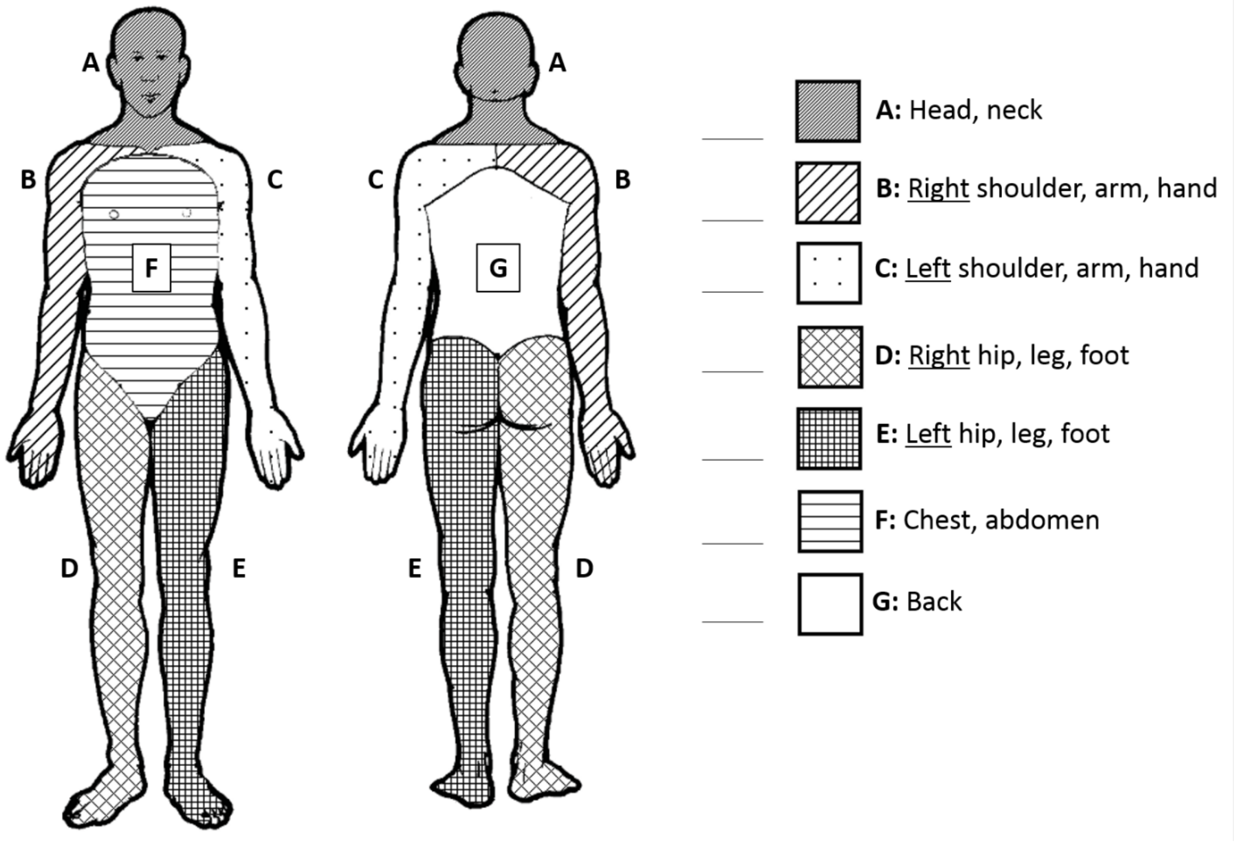


Please read the following list of symptoms. If you have had any of these symptoms for at least three (3) months in the past year, please mark the appropriate box.

Numbness or tingling in arms or legs ☐

Nausea ☐

Sensitivity to odors ☐

Sensitivity to sound ☐
